# Willingness of Nigerian residents to disclose COVID-19 symptoms and take COVID-19 test

**DOI:** 10.1101/2020.10.02.20205914

**Authors:** Victoria Oladoyin, Oluyemi Okunlola, Oluwaseyi Israel, Demilade Ibirongbe, Joy Osifo, Taiwo Obembe, Paulinus Omode, Olugbenga Osunmakinwa

## Abstract

**Background:** An understanding of willingness of people to disclose coronavirus disease 2019 (COVID-19) symptoms and take the COVID-19 test will help provide important insight for motivators towards the self-surveillance and testing strategies recommended by the World Health Organization to curtail and halt the transmission of COVID-19.

**Objectives:** This study assessed willingness to disclose symptoms suggestive of COVID-19 and willingness to take COVID-19 test as well as their predictors.

**Methods:** A cross-sectional online survey of 524 Nigerian adults, aged ≥ 18 years, residing in Nigeria and who had not taken the COVID-19 test was conducted. Information on willingness to disclose COVID-19 symptoms, take COVID-19 test and possible predictors were collected. Data were analysed using descriptive and inferential statistics evaluated at 5% significance level.

**Results:** Mean age of respondents was 35.8 ± 10.7 years and 57.0% were males. Majority (85.8% and 86.2% respectively) were willing to disclose COVID-19 symptoms and take COVID-19 test. Self-risk perception of contracting COVID-19 predicted both willingness to disclose COVID-19 symptoms (aOR=3.236; 95%CI=1.836-5.704) and take COVID-19 test (aOR=3.174; 95%CI=1.570-6.419). Willingness to disclose COVID-19 symptoms (aOR=13.060; 95%CI= 6.253-27.276), knowledge of someone who had taken the test (aOR= 4.106; 95%CI= 1.179-14.299) and thought that it was important for people to know their COVID-19 status (aOR=3.123; 95%CI= 1.516-6.434) also predicted willingness to take COVID-19 test.

**Conclusion:** Nigerians are willing to disclose symptoms suggestive of COVID-19 and take the COVID-19 test. Investment in interventions developed based on the predicting factors will help speed up the finding and testing of suspected COVID-19 cases.

## Introduction

The coronavirus disease 2019 (COVID-19) which started as a localised outbreak in China has since evolved into a global pandemic with countries at different stages of national and subnational outbreak. ^1, 2^ Irrespective of the stage of COVID-19 outbreak experienced by a country, the overarching goal of the global strategy in responding to COVID-19 is for all countries to control the pandemic by slowing down the transmission and reducing mortality associated with COVID-19. ^2^ In the absence of a vaccine or drug to curtail the spread of the disease, some other measures recommended by the World Health Organization (WHO) to help slow down the transmission of COVID-19 with the ultimate aim of reaching and/or maintaining a steady state of low-level or no transmission include finding, testing, isolating and caring for cases and quarantining contacts. ^2^

To identify cases, the WHO recommends that in addition to active case finding in communities, health facilities, and at point of entries, it will be necessary for countries and communities to enable the general population to practice self-surveillance, in which individuals are asked to self-report as a suspected case as soon as they have symptoms or signs and/or if they have contact with a confirmed case. ^2^ Following identification of suspected case, immediate COVID-19 testing is recommended to confirm or rule out infection with COVID-19, in contexts where testing is possible. ^2^

Substantial efforts have been made by the Nigerian Government in implementing self-surveillance and COVID-19 testing. The Nigerian Government, together with partners, Non-Governmental Organizations, the private sector and many other sectors have been encouraging the Nigerian populace, using different media, to get in touch with public health officials via phone calls and WhatsApp messages when they have symptoms suggestive of COVID-19. Also, the Nigerian Government has also been conducting COVID-19 test on people with suspected COVID-19 case definition, hospitalised severe acute respiratory patients and contacts of confirmed patients who are ill. It is however noteworthy to mention that Nigeria is currently in the COVID-19 community transmission phase and to overcome the spread of the disease during this phase, the cases need to be identified very fast by testing and more testing. ^2^

Having highlighted the importance of self-surveillance and testing, success of these strategies will however be dependent on people’s willingness to disclose COVID-19 symptoms and willingness to take the COVID-19 test which are also dependent on several factors such as people’s perception of COVID-19 and their acceptability of COVID-19 testing. ^3^ Fear of the unknown about COVID-19 is another factor that can determine the success of the strategies. With progress of the coronavirus pandemic in Nigeria, false and misleading claims in the media have continued to spread more than the disease itself. False and misleading information in the past have been found to fuel the fear of HIV stigmatization and discrimination when HIV was a pandemic with a resultant lack of willingness to access testing, treatment and care even when these became so widely available for HIV. ^4^ It will not be out of place therefore to assess people’s willingness to disclose their symptoms of COVID-19, their willingness to take COVID-19 test and the factors associated with their decisions if early flattening of the COVID-19 epidemic curve and a halt on COVID-19’s toll on individuals, families, communities and societies in Nigeria and globally are desired. An understanding of the willingness of the community to disclose symptoms and take COVID-19 test will also provide important insight for behavioral change communication for preventive and management measures.

In this study therefore, we assessed the willingness of Nigerians to disclose symptoms suggestive of COVID-19 to designated public health authority and their willingness to take the COVID-19 test. We also determined the predictors of willingness to disclose symptoms suggestive of COVID-19 to designated public health authority, as well as the predictors of willingness to take the COVID-19 test among our study populace.

## Material and methods

### Study area

This study was carried out in Nigeria, the most populated country in Africa. The country’s population is estimated to be about 206 million. ^5^ The first case of COVID-19 in Nigeria was discovered in February, 2020. ^6^ Since then, it has spread to all 36 states in the country as well as the Federal Capital territory. Strategies to control transmission of the disease in the country is in line with that recommended by the WHO and this includes self-surveillance and COVID-19 testing.

### Study design, study population and sampling procedure

In order to adhere with the lock down and physical distancing guidelines as at the time of data collection, this study was conducted using an online cross-sectional study design. The study population comprised of Nigerian adults who were 18 years or older, living in Nigeria as at the time of the survey, had not taken the COVID-19 test and consented to participating in the study.

Assuming a 50% proportion of people willing to take the COVID-19 test (no previous study was found) and a 20% non-response rate, a sample size of 480 was calculated for this study using Leslie Kish’s formula for single proportion. This was however approximated to 500. We used a convenience sampling method to recruit our study respondents by using the researchers’ social networks and other online/internet sources. Overall, 524 respondents consented, filled, and submitted the survey.

### Data collection

This study employed the use of a pre-tested questionnaire scripted on Google form to collect information from the respondents. The questionnaire, which was adapted from the WHO HIV testing, treatment and prevention generic tools for operational research, ^7^ was used to collect information on respondents’ socio-demographics, awareness and source of information about COVID-19, perception about COVID-19, willingness to disclose COVID-19 symptoms and willingness to take COVID-19 test.

### Data analysis

The primary outcome variable for this study was willingness to take COVID-19 test while the secondary outcome variable was willingness to disclose symptoms suggestive of COVID-19.

Data uploaded to the Google form server was downloaded in Microsoft Excel format before it was exported to SPSS software version 22 for analysis. Data of respondents who did not meet the inclusion criteria and those with duplicate data were deleted from the dataset. Frequencies and percentages were generated for categorical variables of interest while age of the respondents was summarized using mean and standard deviation. Inferential statistics was carried out at 5% level of statistical significance using Chi-square test. Thereafter, all socio-demographic variables and other variables that were significant at 10% level on bivariate analysis were taken to the multivariate analysis. Multivariate analysis was done using binary logistic regression to determine the predictors of willingness to disclose symptoms suggestive of COVID-19 and willingness to take COVID-19 test at 5% level of statistical significance.

### Ethical considerations

Ethical approval to conduct the study was obtained from the Ondo State Health Research Ethics Committee with study protocol number OSHREC/28/05/20/268. The purpose of the study was also explained to the participants and written electronic consent was obtained from them before they could take the online survey. Respondents who did not give their written electronic consent were disqualified from continuing the online survey. All data used for the survey were uploaded to Google Drive which was accessible only to the Principal Investigator and the Data Auditor. All study respondents’ data downloaded from the Google Drive were kept on two pass-worded computers which were accessible only to the Principal Investigator and the Data Auditor as well.

## Results

Out of the 524 respondents who consented, filled and submitted the survey, only 485 (92.6%) were retained for further analysis.

About half (54.6%) of the respondents fall within the 30 to 49 years age bracket with a mean age of 35.8 ± 10.7 years. There was slightly (50.7%) more male respondents. Majority (90.7%) of the respondents were Christian and most (95.1%) had completed tertiary education. (Table 1)

**Table 1:** Socio-demographics of respondents

| **Variables** | **Frequency (N = 485)** | **Percentage (%)** |
| --- | --- | --- |
| **Age in years** |  |  |
| 18 – 29 | 162 | 33.4 |
| 30 – 49 | 265 | 54.6 |
| 50 – 64 | 53 | 10.9 |
| 65 and above | 5 | 1.0 |
| **Age in years: Mean ± SD\*** | 35.8 ± 10.7 | |
| **Sex** |  |  |
| Male | 246 | 50.7 |
| Female | 239 | 49.3 |
| **Marital status** |  |  |
| Never married | 189 | 39.0 |
| Currently married | 286 | 59.0 |
| Cohabiting | 1 | 0.2 |
| Separated | 8 | 1.6 |
| Widowed | 1 | 0.2 |
| **Religion** |  |  |
| Christianity | 440 | 90.7 |
| Islam | 38 | 7.8 |
| Traditional | 4 | 0.8 |
| Others | 3 | 0.6 |
| **Ethnic group** |  |  |
| Yoruba | 372 | 76.7 |
| Hausa | 7 | 1.4 |
| Ibo | 44 | 9.1 |
| Others | 62 | 12.8 |
| **Highest level of school completed** |  |  |
| No formal education | 0 | 0.0 |
| Primary | 1 | 0.2 |
| Junior secondary | 0 | 0.0 |
| Senior secondary | 23 | 4.7 |
| Tertiary | 461 | 95.1 |
| **Current occupational status** |  |  |
| Business owners | 68 | 14.0 |
| Employed | 329 | 67.8 |
| Unemployed | 88 | 18.1 |

\*Standard deviation

Most (99.0%) of the respondents agreed that COVID-19 is real, 86.0% disagreed that not everybody is at risk of contracting the disease and 71.4% agreed that they were at risk of contracting the disease. (Table 2)

**Table 2:** Perception about COVID-19

| **Variables**<br>**(N = 485)** | **Strongly**<br>**agree (%)** | **Agree**<br>**(%)** | **Disagree**<br>**(%)** | **Strongly**<br>**disagree**<br>**(%)** |
| --- | --- | --- | --- | --- |
| COVID-19 is real | 364(75.1) | 116(23.9) | 4(0.8) | 1(0.2) |
| COVID-19 is a disease of the rich people only | 6(1.2) | 7(1.4) | 159(32.8) | 313(64.5) |
| Not everybody is at risk of contracting COVID-19 | 22(4.5) | 46(9.5) | 186(38.4) | 231(47.6) |
| I am at risk of contracting COVID-19 | 123(25.4) | 223(46.0) | 94(19.4) | 45(9.3) |
| Since there is no cure for COVID-19, if people have symptoms suggestive of COVID-19, there is no need to disclose their symptoms to public health officials | 19(3.9) | 11(2.3) | 148(30.5) | 307(63.3) |
| Although there is no cure for COVID-19, if people have symptoms suggestive of COVID-19, they should be willing to take the COVID-19 test | 322(66.4) | 150(30.9) | 8(1.6) | 5(1.0) |

Majority of our study respondents were aware of the mode of transmission (96.5%) of COVID-19, its symptoms (99.0%) and how it can be prevented (94.4%). The main sources of information about COVID-19 among our study respondents were television (90.3%) and WhatsApp (81.6%). (Table 3)

**Table 3:** Willingness to disclose COVID-19 symptoms and take a COVID-19 test

| **Willingness to disclose COVID-19 symptoms** | **Yes**<br>**(N=485)** | **%** |
| --- | --- | --- |
| COVID-19 information respondents have heard about* |  |  |
| Transmission | 468 | 96.5 |
| Symptoms | 469 | 96.7 |
| Prevention | 458 | 94.4 |
| Treatment | 249 | 51.3 |
| Others | 15 | 3.1 |
| Source of information about COVID-19* |  |  |
| Television | 438 | 90.3 |
| Radio | 359 | 74.0 |
| Text message (sms) | 386 | 79.6 |
| Health centre/hospital | 196 | 40.4 |
| WhatsApp | 396 | 81.6 |
| Facebook | 290 | 59.8 |
| Twitter | 193 | 39.8 |
| Instagram | 162 | 33.4 |
| Other online (internet) sources | 257 | 53.0 |
| Posters | 174 | 35.9 |
| Flyers | 143 | 29.5 |
| Friends | 266 | 54.8 |
| Community members | 159 | 32.8 |
| Church | 0 | 0 |
| Mosque | 0 | 0 |
| Others | 9 | 1.9 |
| If someone has symptoms suggestive of COVID-19, do you think that person should tell anyone else? | 453 | 93.4 |
| Knowledge about availability of public health hotlines that can be called if a person has symptoms suggestive of COVID-19 | 477 | 98.4 |
| Willingness of respondent to disclose symptoms suggestive of COVID-19 to public health officials via COVID-19 public health hotlines | 416 | 85.8 |
| **Willingness to take COVID-19 test** | **Yes**<br>**(N=485)** | **%** |
| Awareness about COVID-19 test | 480 | 99.0 |
| Ever thought of taking a COVID-19 test | 124 | 25.6 |
| Do you know anyone personally who has been tested for COVID-19? | 90 | 18.6 |
| Do you think it is important for people to know their COVID-19 status? | 397 | 81.9 |
| Willingness of respondent to take COVID-19 test if they have symptoms suggestive of COVID-19 | 418 | 86.2 |
| Willingness of respondent to take COVID-19 test if they do not have symptoms suggestive of COVID-19 | 210 | 43.3 |
| *Multiple response |  |  |

Although almost all (98.4%) of the respondents knew that there were public health hotlines that can be called if a person has symptoms suggestive of COVID-19, a slightly lower number (85.8%) were willing to disclose symptoms suggestive of COVID-19 to public health officials through COVID-19 public health hotlines if they had symptoms suggestive of COVID-19. (Table 3)

Even though 99.0% of the respondents were aware that there was a test for COVID-19, only 18.6% personally knew someone who had taken the COVID-19 test and only 25.6% had ever thought of taking the COVID-19 test. Quite a lot (81.9%) of the respondents however thought it was important for people to know their COVID-19 status and majority (86.2%) of them were willing to take the test if they have symptoms suggestive of the disease. (Table 3)

On bivariate analysis, the perception that not everybody is at risk of contracting COVID-19 and self-perceived risk of contracting COVID-19 by the respondents were both associated with willingness of respondents to disclose symptoms suggestive of COVID-19 to public health officials via COVID-19 public health hotlines. These associations were statistically significant (p=0.018 and p<0.001 respectively). In logistic regression however, the only predictor of willingness to disclose symptoms suggestive of COVID-19 to public health officials via COVID-19 public health hotlines was self-perceived risk of contracting COVID-19 by the respondents. Respondents who agreed that they were at risk of contracting COVID-19 were about three times more likely to disclose their symptoms to public health officials via COVID-19 public health hotlines than those respondents who disagreed (aOR=3.236; 95%CI=1.836-5.704). (Table 4)

**Table 4:**
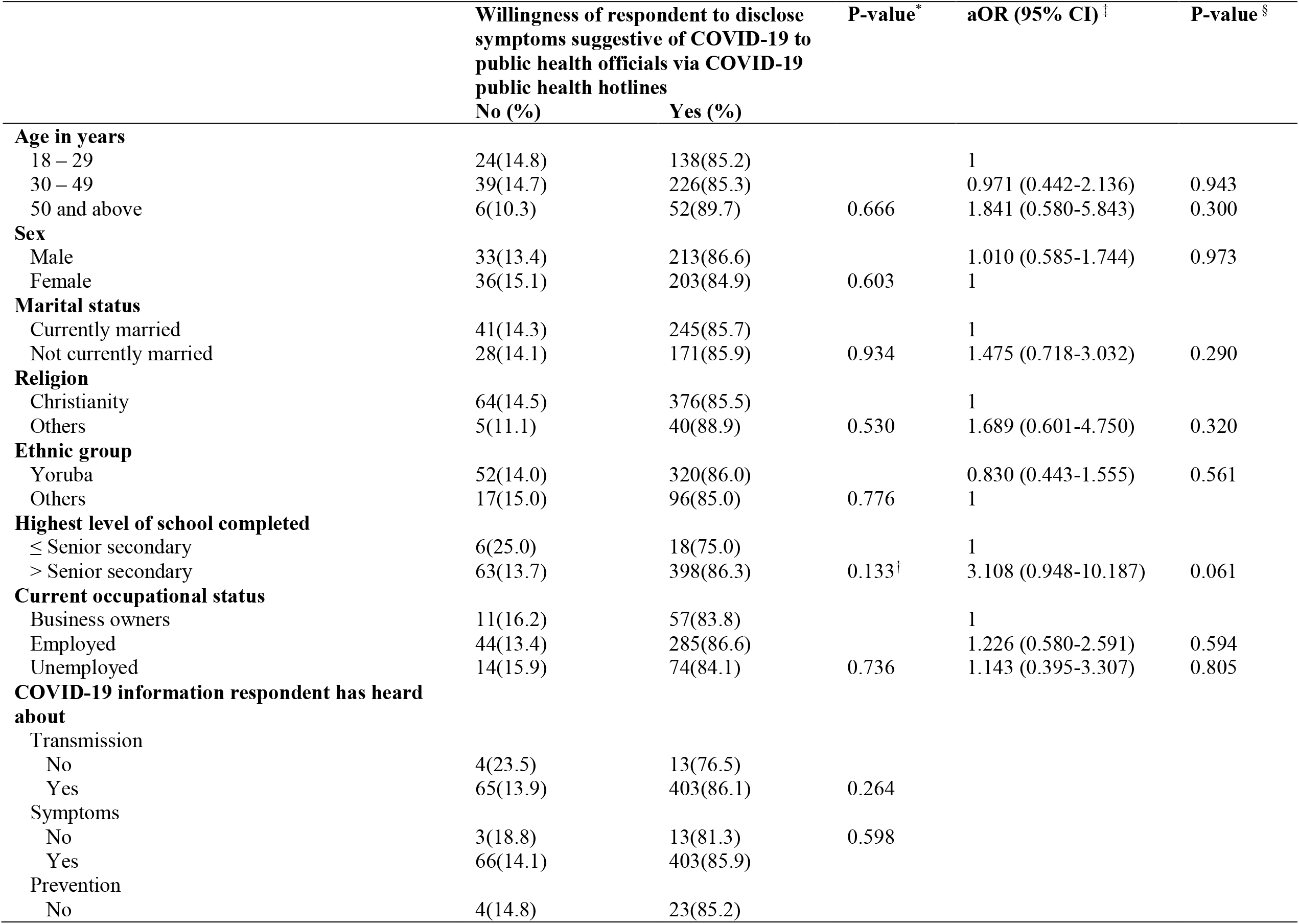
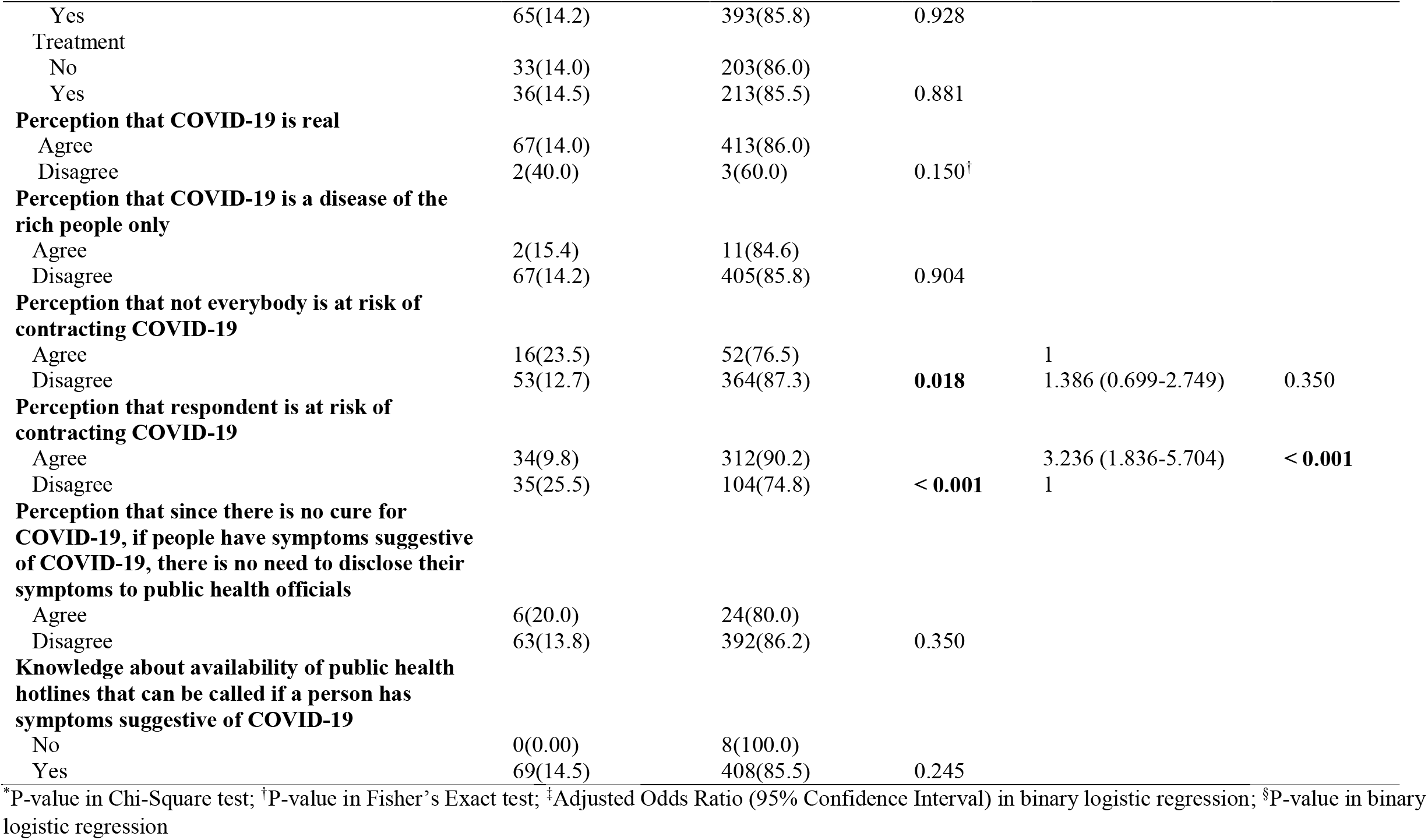
Predictors of willingness to disclose COVID-19 symptoms

Statistically significant associations with willingness to take the COVID-19 test if respondents have symptoms suggestive of the disease on bivariate analysis were perception that COVID-19 is real (p=0.021); self-perceived risk of contracting COVID-19 by the respondents (p<0.001); perception that although there is no cure for COVID-19, if people have symptoms suggestive of COVID-19, they should be willing to take the COVID-19 test (p=0.023); willingness to disclose symptoms suggestive of COVID-19 to public health officials via COVID-19 public health hotlines (p<0.001); thought of taking the COVID-19 test by the respondent (p<0.001); knowing someone who has taken the test (p=0.012); and thought that it was important for people to know their COVID-19 status (p<0.001). (Table 5)

**Table 5:**
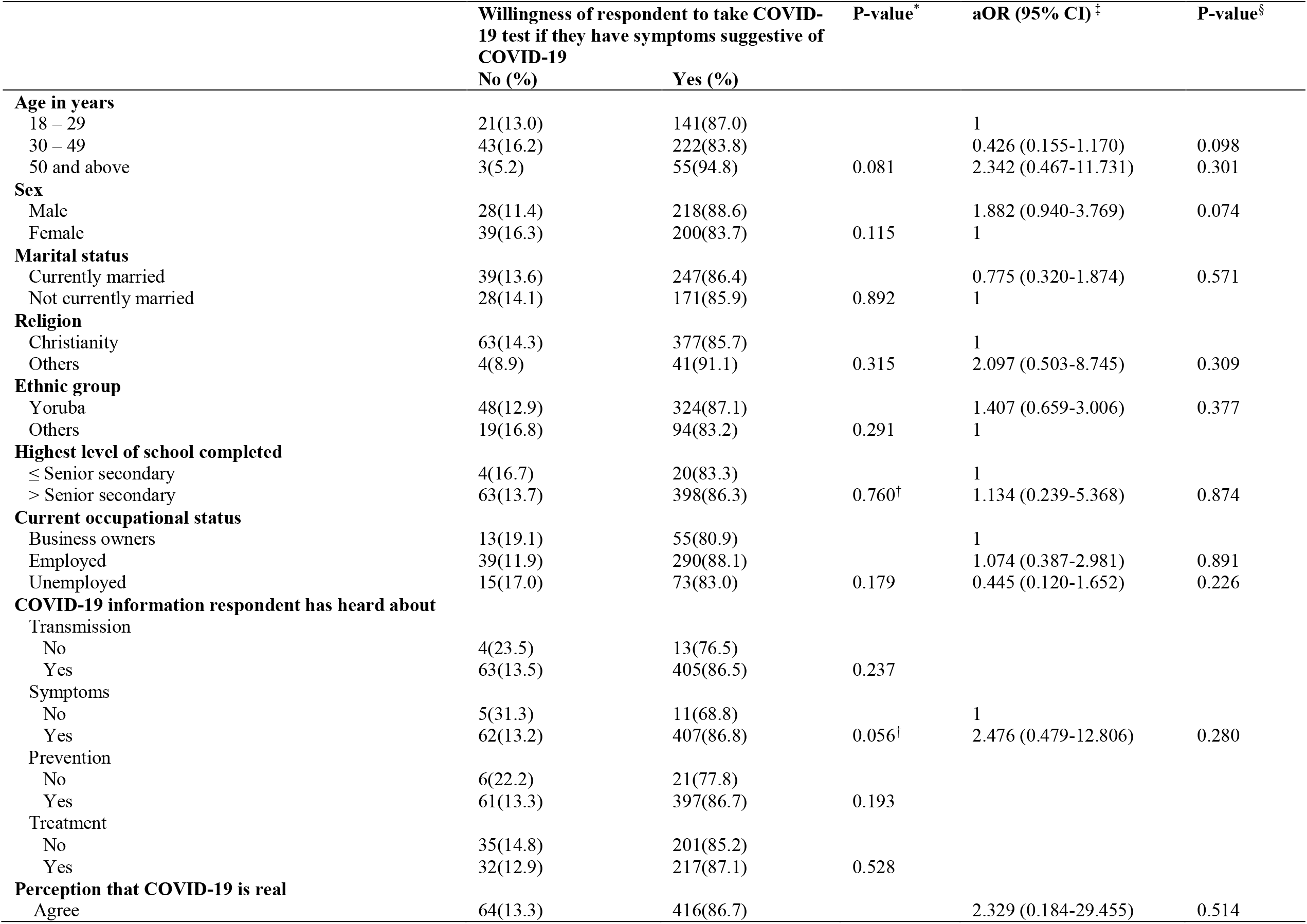
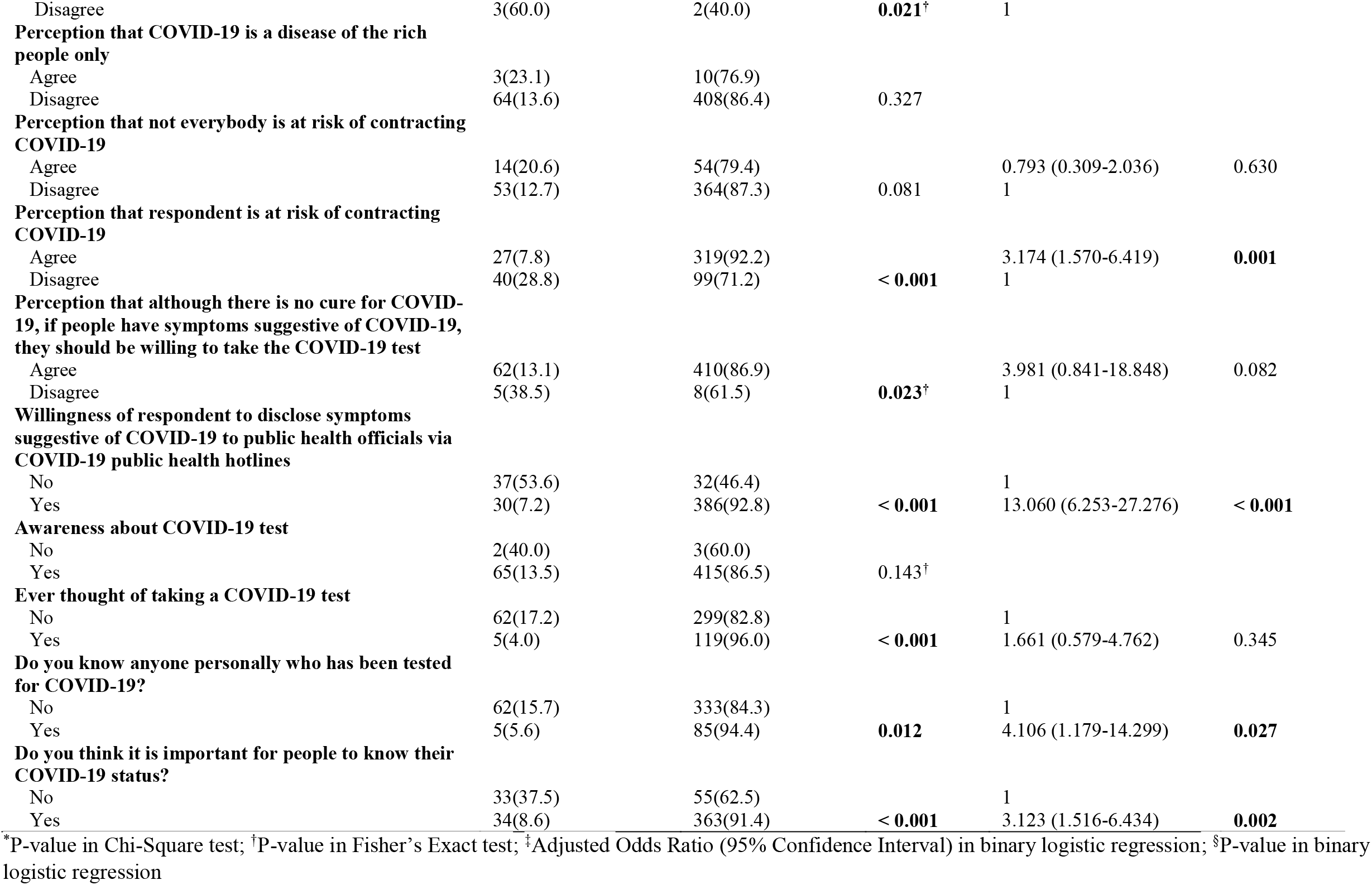
Predictors of willingness to take COVID-19 test

|  | Willingness of respondent to take COVID-19 test if they have symptoms suggestive of COVID-19 |  | P-value* | aOR (95% CI) ‡ | P-value§ |
| --- | --- | --- | --- | --- | --- |
|  | No (%) | Yes (%) |  |  |  |
| **Age in years** |  |  |  |  |  |
| 18 – 29 | 21(13.0) | 141(87.0) |  | 1 |  |
| 30 – 49 | 43(16.2) | 222(83.8) |  | 0.426 (0.155-1.170) | 0.098 |
| 50 and above | 3(5.2) | 55(94.8) | 0.081 | 2.342 (0.467-11.731) | 0.301 |
| **Sex** |  |  |  |  |  |
| Male | 28(11.4) | 218(88.6) |  | 1.882 (0.940-3.769) | 0.074 |
| Female | 39(16.3) | 200(83.7) | 0.115 | 1 |  |
| **Marital status** |  |  |  |  |  |
| Currently married | 39(13.6) | 247(86.4) |  | 0.775 (0.320-1.874) | 0.571 |
| Not currently married | 28(14.1) | 171(85.9) | 0.892 | 1 |  |
| **Religion** |  |  |  |  |  |
| Christianity | 63(14.3) | 377(85.7) |  | 1 |  |
| Others | 4(8.9) | 41(91.1) | 0.315 | 2.097 (0.503-8.745) | 0.309 |
| **Ethnic group** |  |  |  |  |  |
| Yoruba | 48(12.9) | 324(87.1) |  | 1.407 (0.659-3.006) | 0.377 |
| Others | 19(16.8) | 94(83.2) | 0.291 | 1 |  |
| **Highest level of school completed** |  |  |  |  |  |
| ≤ Senior secondary | 4(16.7) | 20(83.3) |  | 1 |  |
| > Senior secondary | 63(13.7) | 398(86.3) | 0.760† | 1.134 (0.239-5.368) | 0.874 |
| **Current occupational status** |  |  |  |  |  |
| Business owners | 13(19.1) | 55(80.9) |  | 1 |  |
| Employed | 39(11.9) | 290(88.1) |  | 1.074 (0.387-2.981) | 0.891 |
| Unemployed | 15(17.0) | 73(83.0) | 0.179 | 0.445 (0.120-1.652) | 0.226 |
| **COVID-19 information respondent has heard about** |  |  |  |  |  |
| Transmission |  |  |  |  |  |
| No | 4(23.5) | 13(76.5) |  |  |  |
| Yes | 63(13.5) | 405(86.5) | 0.237 |  |  |
| Symptoms |  |  |  |  |  |
| No | 5(31.3) | 11(68.8) |  | 1 |  |
| Yes | 62(13.2) | 407(86.8) | 0.056† | 2.476 (0.479-12.806) | 0.280 |
| Prevention |  |  |  |  |  |
| No | 6(22.2) | 21(77.8) |  |  |  |
| Yes | 61(13.3) | 397(86.7) | 0.193 |  |  |
| Treatment |  |  |  |  |  |
| No | 35(14.8) | 201(85.2) |  |  |  |
| Yes | 32(12.9) | 217(87.1) | 0.528 |  |  |
| **Perception that COVID-19 is real** |  |  |  |  |  |
| Agree | 64(13.3) | 416(86.7) |  | 2.329 (0.184-29.455) | 0.514 |
| Disagree | 3(60.0) | 2(40.0) | **0.021<sup>†</sup>** | 1 |  |
| **Perception that COVID-19 is a disease of the rich people only** |  |  |  |  |  |
| Agree | 3(23.1) | 10(76.9) |  |  |  |
| Disagree | 64(13.6) | 408(86.4) | 0.327 |  |  |
| **Perception that not everybody is at risk of contracting COVID-19** |  |  |  |  |  |
| Agree | 14(20.6) | 54(79.4) |  | 0.793 (0.309-2.036) | 0.630 |
| Disagree | 53(12.7) | 364(87.3) | 0.081 | 1 |  |
| **Perception that respondent is at risk of contracting COVID-19** |  |  |  |  |  |
| Agree | 27(7.8) | 319(92.2) |  | 3.174 (1.570-6.419) | **0.001** |
| Disagree | 40(28.8) | 99(71.2) | **< 0.001** | 1 |  |
| **Perception that although there is no cure for COVID-19, if people have symptoms suggestive of COVID-19, they should be willing to take the COVID-19 test** |  |  |  |  |  |
| Agree | 62(13.1) | 410(86.9) |  | 3.981 (0.841-18.848) | 0.082 |
| Disagree | 5(38.5) | 8(61.5) | **0.023<sup>†</sup>** | 1 |  |
| **Willingness of respondent to disclose symptoms suggestive of COVID-19 to public health officials via COVID-19 public health hotlines** |  |  |  |  |  |
| No | 37(53.6) | 32(46.4) |  | 1 |  |
| Yes | 30(7.2) | 386(92.8) | **< 0.001** | 13.060 (6.253-27.276) | **< 0.001** |
| **Awareness about COVID-19 test** |  |  |  |  |  |
| No | 2(40.0) | 3(60.0) |  |  |  |
| Yes | 65(13.5) | 415(86.5) | 0.143<sup>†</sup> |  |  |
| **Ever thought of taking a COVID-19 test** |  |  |  |  |  |
| No | 62(17.2) | 299(82.8) |  | 1 |  |
| Yes | 5(4.0) | 119(96.0) | **< 0.001** | 1.661 (0.579-4.762) | 0.345 |
| **Do you know anyone personally who has been tested for COVID-19?** |  |  |  |  |  |
| No | 62(15.7) | 333(84.3) |  | 1 |  |
| Yes | 5(5.6) | 85(94.4) | **0.012** | 4.106 (1.179-14.299) | **0.027** |
| **Do you think it is important for people to know their COVID-19 status?** |  |  |  |  |  |
| No | 33(37.5) | 55(62.5) |  | 1 |  |
| Yes | 34(8.6) | 363(91.4) | **< 0.001** | 3.123 (1.516-6.434) | **0.002** |

\*P-value in Chi-Square test; <sup>†</sup>P-value in Fisher's Exact test; <sup>‡</sup>Adjusted Odds Ratio (95% Confidence Interval) in binary logistic regression; <sup>§</sup>P-value in binary logistic regression

The predictors of willingness to take the COVID-19 test if respondents have symptoms suggestive of the disease in binary logistic regression were self-perceived risk of contracting COVID-19 by the respondents, willingness to disclose symptoms suggestive of COVID-19 to public health officials via COVID-19 public health hotlines, knowing someone who has taken the test, and thought that it was important for people to know their COVID-19 status. Respondents who perceived themselves as being at risk of contracting COVID-19 were about three times more willing to take the COVID-19 test if they have symptoms suggestive of the disease (aOR=3.174; 95%CI=1.570-6.419) than those who did not perceive themselves of being at risk of contracting the disease. Respondents who were willing to disclose symptoms suggestive of COVID-19 to public health officials were thirteen times more willing to take the COVID-19 test if they have symptoms suggestive of the disease (aOR=13.060; 95%CI= 6.253-27.276) compared to those who were not willing to disclose symptoms suggestive of COVID-19. Respondents who personally knew someone who had taken the COVID-19 test were about four times more willing to take the COVID-19 test if they have symptoms suggestive of the disease (aOR= 4.106; 95%CI= 1.179-14.299) when compared with a respondent who did not know anyone personally. Respondents who thought that it was important for people to know their COVID-19 status were three times more willing to take the COVID-19 test if they had symptoms suggestive of the disease (aOR=3.123; 95%CI= 1.516-6.434) compared to those who did not think it was important for people to know their status. (Table 5)

## Discussion

Since the beginning of 2020, COVID-19 has caused considerable morbidity and mortality worldwide, and has become a priority of the global society. Finding and testing of suspected cases is one of the WHO recommended strategies for stopping the spread of the disease. ^2^ In this study, we examined the willingness and predictors of disclosure of symptoms suggestive of COVID-19 as well as the willingness and predictors of taking a COVID-19 test.

Our study revealed a high willingness to practice self-surveillance by the study participants if they had symptoms suggestive of COVID-19 by disclosing their symptoms to public health officials using the available media. They were also willing to take the COVID-19 test as long as they had symptoms suggestive of COVID-19. A self-perceived risk of contracting COVID-19 was a common predictor of willingness to disclose COVID-19 symptoms and willingness to take the COVID-19 test among our study respondents.

To the best of our knowledge, documented evidence on willingness to disclose COVID-19 symptoms is scarce. The starting point in disease surveillance and notification, which has been recognized as an effective strategy for the prevention and control of diseases, especially those diseases which are epidemic prone, is identification or detection of cases. ^8-10^ In the case of COVID-19, WHO recommends that in addition to active case finding in communities, health facilities, and at points of entries, the practice of self-surveillance will be an added advantage in identifying the cases of COVID-19. ^2^ The high willingness to self-report symptoms suggestive of COVID-19 to designated public health authority among our study respondents is therefore quite encouraging as this will help identify and investigate as many who have symptoms suggestive of COVID-19 and ultimately help reduce the spread. Our result should however be interpreted with caution because even though we tried to control for socio-demographic confounding variables, most of our respondents were drawn from the researchers’ social network and this may not be representative of our target population. More representative quantitative surveys on willingness to disclose symptoms suggestive of COVID-19 and future similar pandemics is encouraged, especially now that the lock-down in the country has been eased and gradual re-opening of various sectors is being implemented. More robust qualitative studies are also encouraged as this will help identify socio-cultural issues that may hinder disclosure of symptoms suggestive of COVID-19 and how to tackle them.

From our study, a self-perceived risk of contracting the COVID-19 is a possible explanation for the willingness to disclose COVID-19 symptoms. The health belief model predicts that individuals with low self-perceived risk of becoming sick are more likely to engage in unhealthy or risky behaviours/actions and vice versa. ^11^ As such, the finding of individuals with self-perceived risk of contracting COVID-19 in this study with a higher odds to willingly engage in healthy actions, which in this case is disclosure of symptoms suggestive of COVID-19 to public health officials, is not surprising. The implication of this is that, in addition to other information on COVID-19 such as the modes of transmission, symptoms and prevention which are communicated via television, WhatsApp, text messages and radio to the general populace as attested by our study participants and other studies, ^12, 13^ information on risk assessment should also be communicated. This may help improve self-surveillance especially among people with moderate to high risk of contracting COVID-19.

Although our study finding showed that a high proportion of our study respondents were willing to take the COVID-19 test as long as they had symptoms suggestive of the disease, the proportion of people who were willing to take the test decreased from 86.2% if they had symptoms to 43.3% if they do not have symptoms. This finding is corroborated by the findings of Siegler and colleagues in their study on willingness to seek diagnostic test for Severe Acute Respiratory Syndrome Coronavirus 2 (SARS-CoV-2) with home, drive-through, and clinic-based specimen collection locations. ^3^ Siegler and colleagues showed that persons experiencing COVID-19 symptoms had higher mean Likert scale values with respect to willingness to seek diagnostic test for SARS-CoV-2 for each testing modality location studied compared to persons not experiencing symptoms at the time of the study. It is however important to note that current research has demonstrated the existence of asymptomatic and pre-symptomatic COVID-19 cases who are potential transmitters of the disease. ^14-17^ In the light of this, even though it may not be possible to test everyone, balanced information on presence of symptoms and engagement in high risk activities should be provided as criteria for testing by the various media used for disseminating COVID-19 information so that people can make themselves available for testing whenever the need arises even if they do not show symptoms. This will help with early detection and isolation of the true cases and thus breaking the chain of transmission.

The health belief model suggests that people’s belief about health problems, perceived benefits of action, barriers to action, and self-efficacy in the presence of a cue to action explain engagement or lack of engagement in health promoting behaviors. ^11^ The explanatory variables for the willingness to take the COVID-19 test among our study respondents if they had symptoms suggestive of COVID-19 were therefore not surprising as these can be interpreted using some of the constructs of the health belief model such as perceived susceptibility, perceived benefits and cues to action. The implication of our findings is that interventions based on the health belief model can be developed by targeting these predictors as a change tool to increasing the willingness of individuals to take the COVID-19 test, if the need arises. So for example, perceived susceptibility to COVID-19 can be increased by providing more information and education about how individuals can assess their own risk of contracting COVID-19 and the likelihood of contracting COVID-19 based on their risk category. Perceived benefit of taking the COVID-19 test can be increased by providing more information and education on reasons why it is important for people to know their COVID-19 status. Such reasons include, if people know they are COVID-19 positive, they can take appropriate measures to reduce the probability of infecting others and also the fact that the data provided by widespread COVID-19 testing will better inform the social distancing policies. ^18^ Furthermore, interventions based on the health belief model may provide cues to action by bringing up respected opinion leaders, community influencers and volunteers to speak about their COVID-19 experiences using various platforms in order to encourage people to take the COVID-19 test.

A limitation of this study, which must be acknowledged, is the non-representativeness of the study population as most of our respondents were within the researchers’ social network. Despite this limitation, our study was able to add to the body of knowledge on COVID-19.

## Conclusion

Nigerians are willing to disclose symptoms suggestive of COVID-19 and take the COVID-19 test. This willingness increases when there is a perceived susceptibility of contracting the disease. In addition, information about perceived benefits and presence of cues to action increases the chances of taking the COVID-19 test among Nigerians. Investment in interventions targeted at these predictors will help speed up the finding and testing of suspected COVID-19 cases as recommended by WHO and this will ultimately lead to early halting of the spread of the disease.

## Data Availability

All data referred to in the manuscript are available upon request

## Acknowledgements

We thank all the respondents who participated in this study. We also acknowledge the contributions of all who assisted with data collection.

